# Identification of high-risk COVID-19 patients using machine learning

**DOI:** 10.1101/2021.02.10.21251510

**Authors:** Mario A. Quiroz-Juárez, Armando Torres-Gómez, Irma Hoyo-Ulloa, Roberto de J. León-Montiel, Alfred B. U’Ren

## Abstract

The current COVID-19 public health crisis, caused by SARSCoV-2 (severe acute respiratory syndrome coronavirus 2), has produced a devastating toll both in terms of human life loss and economic disruption. In this paper we present a machine-learning algorithm capable of identifying whether a given patient (actually infected or suspected to be infected) is more likely to survive than to die, or vice-versa. We train this algorithm with historical data, including medical history, demographic data, as well as COVID-19-related information. This is extracted from a database of confirmed and suspected COVID-19 infections in Mexico, constituting the official COVID-19 data compiled and made publicly available by the Mexican Federal Government. We demonstrate that the proposed method can detect high-risk patients with high accuracy, in each of four identified treatment stages, thus improving hospital capacity planning and timely treatment. Furthermore, we show that our method can be extended to provide optimal estimators for hypothesis-testing techniques commonly-used in biological and medical statistics. We believe that our work could be of use in the context of the current pandemic in assisting medical professionals with real-time assessments so as to determine health care priorities.

## I. NTRODUCTION

Coronavirus infectious disease (COVID-19) is a recently discovered illness caused by severe acute respiratory syndrome coronavirus 2 (SARS-CoV-2). As of the first week of february 2021, over 106 million SARS-CoV-2 infections and over 2.3 million deaths have been registered worldwide, in the worst pandemic to afflict humanity since the so-called Spanish flu of 1918, which has overwhelmed the world’s health care systems and caused severe economic disruption.

As a response to this international public health crisis, scientists and clinicians have made enormous efforts in the last few months to generate new knowledge and to develop technological tools that may help in combatting this infectious disease and mitigate its effects. Some of these efforts include the development of drugs and vaccines [1–4], the construction of epidemiological models to forecast the dynamics of disease spreading in the population [5–8], the development of mobile-device applications for tracking infected patients and new cases [9–11], and the development of strategies and the application of new technologies to manage the resources and capacities in hospitals [12–14].

An emergency non-pharmaceutical prevention measure adopted in many countries has been the reduction or suspension of non-essential activities so as to reduce both the rate of new infections [15] and the risk of exceeding hospital capacities. Undoubtedly, the ability to rapidly identify high-risk patients and/or correctly assign health care priorities is critical, in the first case so as to improve hospital capacity planning and in the second case for providing timely treatment for patients [16]. In this regard, artificial intelligence methods have been recognized as a powerful and promising technology that can help not only in the identification of the fatality risk of a given patient seeking medical attention [17, 18], but also for the diagnosis process [19–22], prediction of disease spreading dynamics [23–27], and tracking of infected patients as well as likely future patients [28].

Machine learning is a branch of the artificial intelligence field which seeks to endow computers with a “learning capacity” using well-defined algorithms, to improve performance or make accurate predictions. Typically, these algorithms learn from past information available, introduced in the form of labeled training sets. Of course, the quality and size of the data-sets used are crucial in ensuring the adequate performance of the algorithm [29, 30]. During the course of the current pandemic machine learning has been used to develop different algorithms that seek to identify, at an early stage, patients who are likely to become infected. These approaches make predictions relying on basic patient information, clinical symptoms [31–33], as well as travel history [34] and discharge time of hospitalized patients [16]. Some other efforts focus on identifying patients requiring specialized care, namely hospitalization and/or specialized care units [35–37], or patients at a higher fatality risk [38, 39].

In this work, we introduce a machine-learning algorithm that effectively identifies high-risk patients among those that may have been exposed to the SARS-CoV-2 virus. Our method employs a supervised artificial neural network which predicts whether a given patient belongs to one of two classes: *class 1*, which represents those patients who are more likely to survive than to die, and *class 2* which represents those patients who are more likely to die than to survive. In order to achieve this classification, we rely on a database with information about past infections (along with suspected infections), from which we extract a 28-element characteristics vector for each patient. The characteristics include information about comorbidities, patient demographic data, as well as recent COVID-19-related medical information. In our algorithm, we apply our rapid identification of patients (belonging either to class 1 or class 2) at any of four clearly-defined stages of the treatment process, ranging from stage 1 at which a patient first becomes ill and seeks medical attention, to stage 4 at which not only is the patient hospitalized but requires specialized attention. We have trained a neural network for each of the four stages, with data corresponding to a characteristics subset, with later stages having access to a larger fraction of the 28 characteristics as they become known during the treatment process. Our algorithm is able to classify patients with high accuracy at each of the four stages, with the accuracy value increasing with the progression from one stage to the next. Furthermore, we demonstrate that our algorithm can provide an optimal estimator for hypothesis-testing techniques commonly-used in biological and medical statistics. This creates a bridge between machine-learning algorithms and clinical medicine that allows for the introduction of a series of novel strategies, with applications in different clinical scenarios, in a familiar language for clinicians. We believe that this technology can be a powerful tool for medical resource allocation and hospital capacity planning, by making correct, real-time assessments of mortality risks given the highly specific characteristics of each particular patient.

## II. DATA

Our studies presented in this paper are based on a publicly-available database of COVID-19 patients from the Mexican Federal Government. This database, which includes all confirmed and suspected COVID-19 cases reported in Mexico, is available in the ‘Statistical Morbidity Yearbooks’ (Anuarios Estadísticos de Morbilidad) published by the General Council of Epidemiology (Dirección General de Epidemiología), part of the Health Ministry (Secretaría de Salud), Mexican Federal Government [40]. For the period from March to January 2020, this database contains a historical record of 4,700,464 patients who have received medical attention at both public and private medical facilities^1^ in all 32 states, 215,301 of which correspond to confirmed deaths, and 4,485,163 to recovered patients. Note that the data is collected through a form filled by each patient during the admission process at the emergency room, clinic, clinical laboratory, or hospital.

The database includes 28 characteristics for each patient, which can be grouped into three categories: 1) past medical history, 2) demographic data, and 3) information related to the COVID-19 episode. Category 1 (medical history) includes comorbidity information, specifically: diabetes, chronic obstructive pulmonary disease (COPD), asthma, use of immunosuppressive drugs, hypertension, cardiovascular disease, obesity, chronic renal disease, asthma, other chronic illnesses, smoking history, and pregnancy. Category 2 (demographic data) includes gender, age, state of birth, state of residence, whether the patient in question self-identifies as indigenous and/or speaks an indigenous language, is a migrant, or a foreigner. Category 3 (recent medical information) is subdivided into category 3a, which corresponds to those characteristics which may be known by a patient at the point of first becoming ill and receiving medical attention, and category 3b, which corresponds to those characteristics which may become known during the course of medical treatment. Category 3a includes the type of medical facility where the patient is being treated^2^, the state where the medical facility is located, the number of days elapsed from the date of symptom onset to the beginning of treatment, and exposure to confirmed COVID-19 patients. Category 3b includes the COVID-19 status, COVID-19-related pneumonia, hospital and intensive care unit (ICU) admission, as well as the need for mechanical ventilation. COVID-19 status identifies confirmed COVID-19 cases through: i) a positive real-time reverse transcription-polymerase chain reaction (RT-PCR) test or a positive COVID-19 antigen test, ii) clinical-epidemiological association, or iii) designation by a special health committee, iv) patients with a negative RT-PCR result or a negative antigen test, v) patients with an invalid laboratory result, vi) patients whose test was not processed, as well as vii) suspected COVID-19 patients who are awaiting a laboratory result.

Note that a fourth category of characteristics, which could exhibit a high predictive power as part of our patient classification algorithm, would include information about current symptoms. Such information is unfortunately not currently available in the public database at our disposal.

Table I summarizes the 28 characteristics and the three categories discussed above. Note that we have displayed in italics 7 characteristics which we have determined to lack a sufficient predictive power and/or yield inconsistent results (as derived from too small a population with the characteristic in question). From this point onward, the characteristics vector employed throughout the manuscript refers to the resulting shortened 21-element vector.

**TABLE I.**
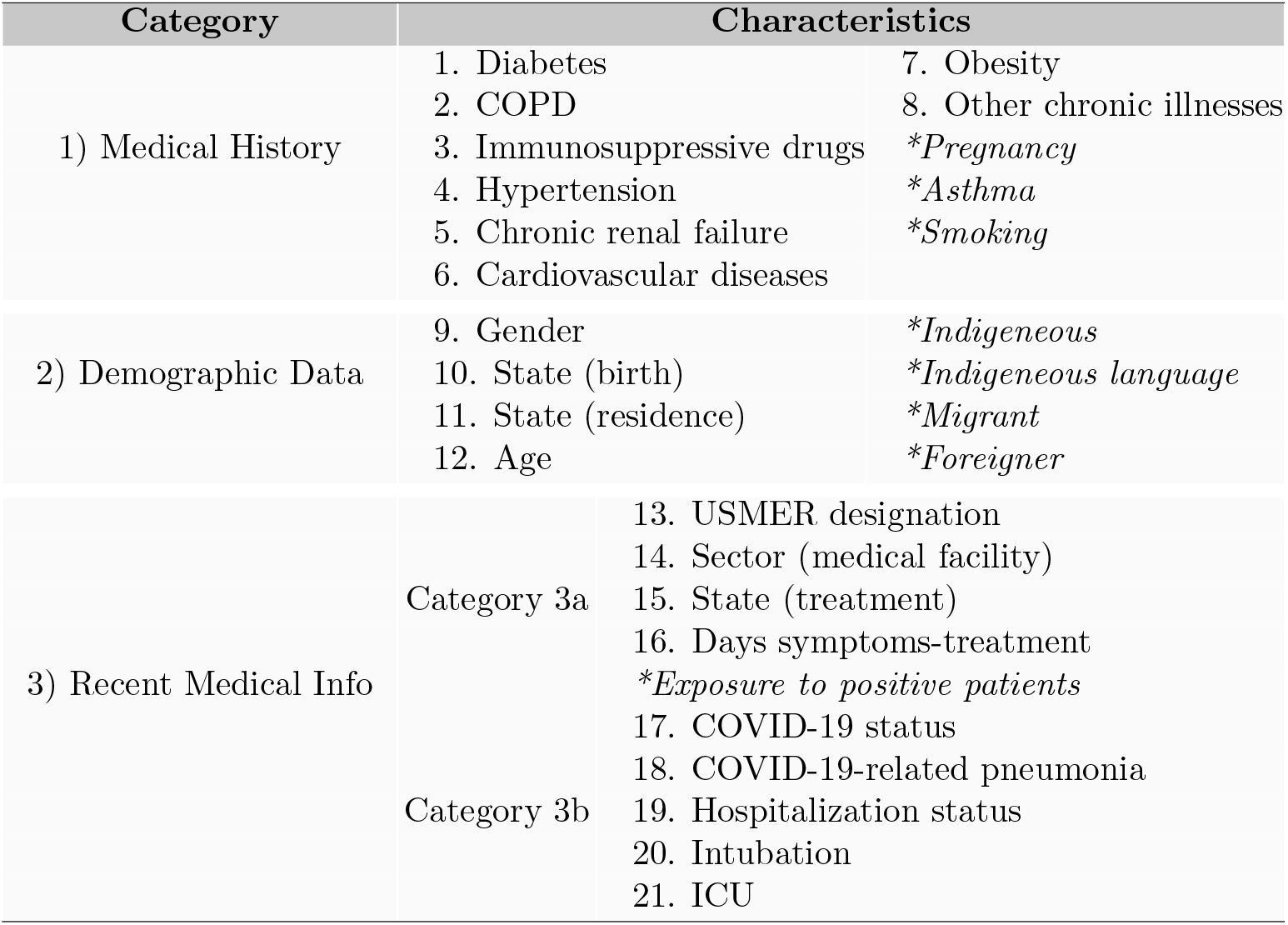
Classification of characteristics. Characteristics shown in italic font do not exhibit a sufficient predictive power and are therefore not included in our subsequent analysis.

## III. CONVENTIONAL TECHNIQUES IN BIOSTATISTICS

In biostatistics, Bayesian inference, hypothesis testing, variance analysis, and regression techniques are methods extensively employed for statistical evaluation of medical data [41, 42]. Along these lines, we have directly applied the hypothesis testing method [43] in order to attempt to discriminate between our two classes of patients from the collected data so as to establish a critical bound which may help us to identify the mortality risk for incoming COVID-19 patients. With this goal in mind, we use the first four central moments as estimators. Figure 1 shows the normalized statistical distributions obtained for class 2, i.e. deceased patients (red), and class 1, i.e. recovered patients (blue), using the moments: (a) mean, (b) variance, (c) skewness, and (d) kurtosis. The central moments were computed from the 21 elements in the characteristics vector. Note that the resulting distributions for class 1 / class 2 patients exhibit a large degree of overlap. This implies that the desired classification of patients through the determination of a critical bound is not possible using these estimators.

**FIG. 1.**
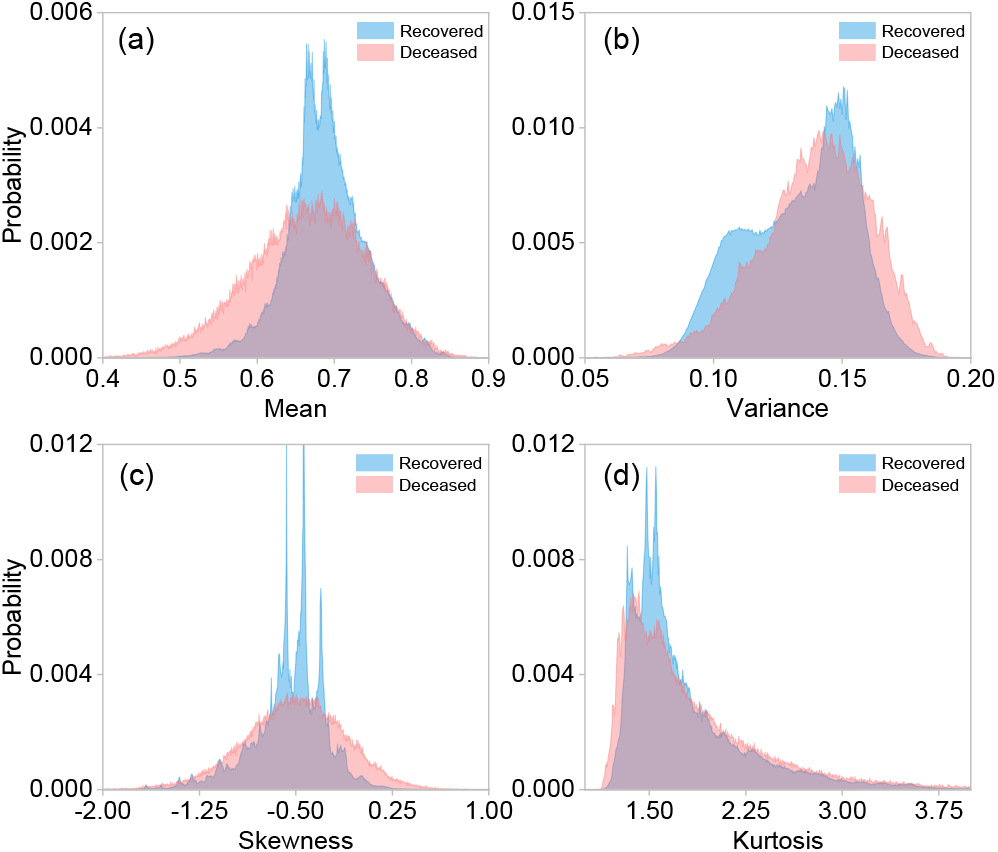
Normalized distributions for different central moments: (a) mean, (b) variance, (c) skewness, and (d) kurtosis, for class 1 (recovered patients), shown in blue and for class 2 (deceased patients), shown in red. These central moments were computed from the 21 elements of the characteristics vector for each patient. These distributions were plotted using the entire dataset with 4, 700, 464 observations.

Bayesian inference constitutes an alternative to frequentist methods, the latter which provide predictions based on the relative frequencies associated with particular events within a large number of trials. Applied to our particular case, Bayesian inference would compute a posterior probability *P* (*C*_*j*_| x), where *C*_*j*_ denotes the relevant classes (recovered, *j* = 1, or deceased, *j* = 2), and x represents the characteristics vector, resulting from a prior probability *P* (*C*_*j*_) and the so-called likelihood function *P* (x |*C*_*j*_) through Bayes theorem [44], *P* (*C*_*j*_| x) = *P* (x |*C*_*j*_)*P* (*C*_*j*_)*/P* (x). Note that in order to determine the likelihood function, *P* (x *C*_*j*_), a model is required in order to estimate the probability of observing a set of characteristics x given a known class *C*_*j*_. Although in principle a viable likelihood function could be derived from the sample data, this method tends to fail because the variances in parameter calibration can be rather large along certain directions of the parameter space [6], linked to the large degree of overlap between the distributions for both classes, which has already been discussed. This implies that in our case Bayesian inference does not provide a functional approach for the discrimination between class 1 (recovered) and class 2 (deceased) patients.

The conventional statistical tools and techniques used in clinical medicine are static processes that are consistent and unchanged. In contrast, machine learning offers a dynamic tool that learns and modifies itself as the learning process develops, producing a more robust tool to make predictions. In this context, it has been shown that artificial intelligence methods provide a novel and promising approach for classification and pattern recognition. In what follows, we design, train, and test neural networks in order to identify individual patients as belonging to either class 1 or class 2, relying on historical data, collected from previous patients.

## IV. NEURAL NETWORK

Machine learning is a method of data analysis that endows computer algorithms with the capacity to “learn” from a known data-set (which includes defining characteristics and an outcome for each observation), so as to produce a prediction about the outcome given a specific choice of characteristics. Of course, the quality and size of the training data set are crucial in determining the resulting performance of the algorithm. In the following we demonstrate the use of neural networks trained with the data-set described in Section II. Our neural networks are then used to predict whether a given patient (not included in the data-set used for training) belongs to one of two classes: class 1, which represents those patients who are more likely to survive than to die, and class 2 which represents, conversely, those patients who are more likely to die than to survive.

The proposed machine learning algorithm is based on a multilayer feed-forward network with two sigmoid neurons in the single hidden layer and two softmax neurons in the output layer. The hidden layer is indicated by a green, dashed-line rectangle in Fig. 2, whereas the output layer is indicated by an orange rectangle. The network’s output represents a probability distribution over the two output classes, which can be interpreted as the survival and mortality probabilities [45, 46]. Figure 2 shows the architecture of the neural networks which we have implemented. The blue lines represent the connections between neurons, each characterized by a synaptic weight. Note that in appendix B we provide plots constituting a detailed report of the dominant synaptic weights for each of the two neurons in the hidden layer.

**FIG. 2.**
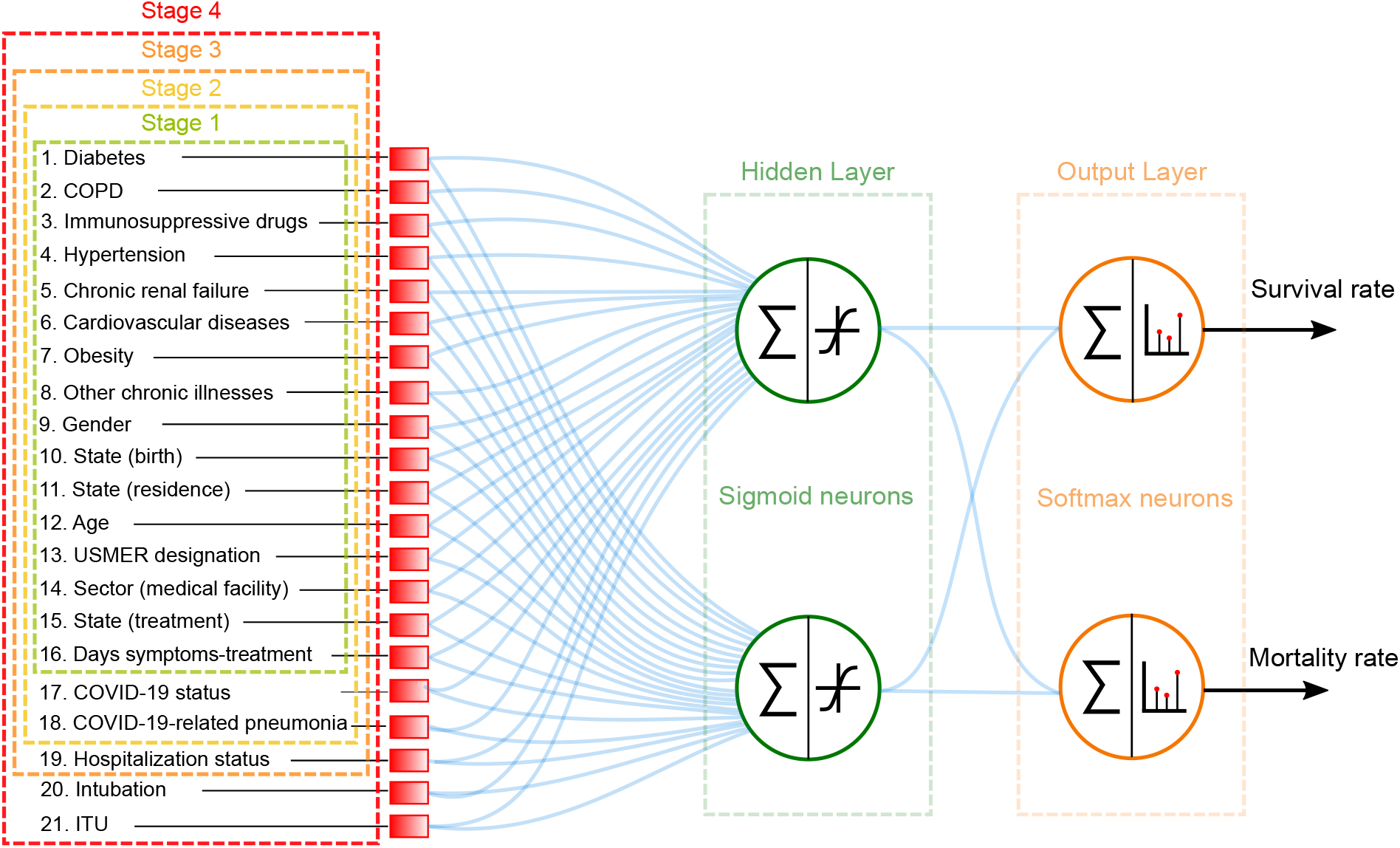
Architecture of the multilayer feed-forward neural network employed in order to classify partients into class 1 (recovered) and class 2 (deceased).

We have designed and trained four separate neural networks, each corresponding to a distinct stage of the treatment process. Patients in stage 1 are those who are in the process of receiving initial medical assessment and care. For these patients, we can assign data for characteristics in categories 1,2, and 3a (see section II and table I). Patients in stage 2 are those who as part of their evaluation already have a known COVID-19 status, and may already have contracted COVID-19-related pneumonia. Therefore, the COVID-19 status and pneumonia characteristics are added to the characteristics vector. Patients in stage 3 are those for whom a decision has been reached as to whether admit into a hospital or send back home. Therefore, the hospitalization status characteristic is added to the characteristics vector. Patients in stage 4 are those who in addition to being hospitalized, have either been intubated or admitted into an intensive care unit. Therefore, the intubation and ICU characteristics are added to the characteristics vector. Note that in Fig. 2, the dashed-colored rectangles indicate the characteristics available in each of the four stages of treatment.

We train our neural networks with the scaled conjugate gradient back-propagation algorithm, while the performance is quantified through the cross-entropy. It is important to point out that, in order to guarantee an unbiased patient classification, we have used a balanced dataset of 430,602 observations, with one half of the observations (215,301) representing all known deceased patients in the database, and the other half (also 215,301) randomly selected from those patients who recovered. This resulting balanced data-set is then divided into three sub-datasets: i) The first one, containing 70% of the observations, for training of the neural networks, ii) a second one, containing 15% of the observations, for validation, and iii) a third one, containing another 15% of the observations, for testing.

In order to assess the performance of our neural networks, we define the specificity or true negative rate, *TNR*, as the share of true negatives (*TN*) - i.e. true recoveries - to the sum of true negatives and false positives (*FP*). We also define the sensitivity or true positive rate, *TPR*, as the share of true positives (*TP*) - i.e. true deaths - to the sum of true positives and false negatives (*FN*). In addition to the specificity and the sensitivity, we also define the accuracy (*ACC*), as follows

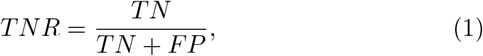

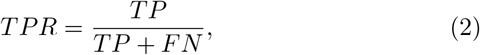

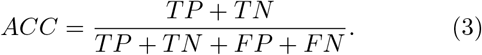

Once training has been completed, our neural networks can predict with high accuracy whether a given patient with known characteristics belongs in class 1 (more likely to survive), or in class 2 (more likely to die). Across all four stages, our machine learning algorithm exhibits a specificity greater than 82%, a sensitivity greater than 86%, and an accuracy greater than 84%. In general, since more information becomes available for successive stages, it is only natural that the specificity, sensibility, and accuracy all tend to improve when progressing from one stage to the next. The accuracy, specificity, and sensitivity reach values of 93.5%, 90.9%, and 96.1%, respectively, at stage 4. The three quantities for each of the four stages are shown in Table II.

**TABLE II.** Accuracy, specificity and sensitivity obtained by the neural networks for each of the four stages.

| Stage | Accuracy | Specificity | Sensitivity |
| --- | --- | --- | --- |
| 1 | 84.3% | 82.4% | 86.3% |
| 2 | 90.5% | 89.1% | 91.9% |
| 3 | 93.1% | 90.8% | 95.5% |
| 4 | 93.5% | 90.9% | 96.1% |

We would like to remark that, given the user-friendly nature of our algorithms, it would be straightforward to port them to hand-held electronic devices, such as mobile phones and tablets [47]. This could facilitate the deployment of this technology across clinics and hospitals, for the real-time identification of high-risk patients. More importantly, it could help identifying the effects of all individual characteristics of each patient in question. Note that, the computed survival probability at each of the four treatment stages could serve as a numerical scale to aid in the allocation of medical resources and the management of hospital capacity.

We would like to stress that while the data at our disposal, with which our algorithms were trained, pertains to Mexico, our algorithm could be easily adapted to data from other countries. Note also that we expect that if we had access to current symptoms data for each patient, the predictions provided by our neural networks would exhibit a considerably enhanced accuracy. We are of course interested in running our algorithm with other data-sets which may become available to us, as our contribution to curbing the current health crisis.

## V. HYBRID MACHINE-LEARNING-ASSISTED FREQUENTIST HYPOTHESIS-TESTING METHOD

As discusses in Section III, none of the first four central moments as calculated from the 21 element of the characteristics vector, provide a viable estimator for the hypothesis testing method. In general, the determination of a viable estimator is a challenging task. Here we propose the use of the softmax outputs of our neural networks as estimators, in other words we let the training of our neural networks accomplish the non-trivial task of determining a highly-optimized estimator. This becomes a hybrid technique which exploits both machine learning and standard hypothesis testing.

In figure 3, we show the resulting distributions of the outputs of each of the four neurons in our neural networks, for those patients known to have died (red), and for those known to have recovered (blue). While the two neurons (particulary the first one) in the internal layer can already do a reasonable job at discriminating between classes 1 and 2, the two neurons in the outer layer accomplish this classification remarkaby well.

**FIG. 3.**
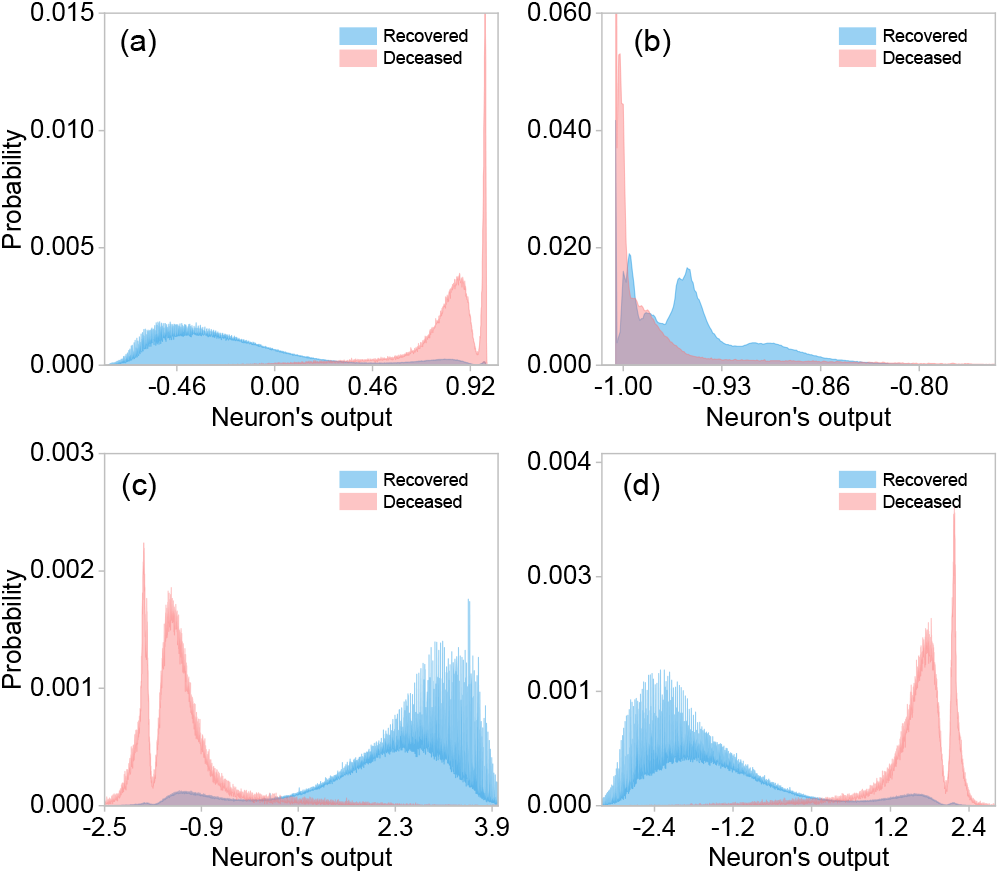
Statistical distributions for deceased (red) and recovered (blue) patients built from the outputs of: (a) neuron-1 in layer 1, (b) neuron-2 in layer 1, (c) neuron-1 in layer 2 and (d) neuron-2 in layer 2.

In order to evaluate the proposed hybrid method, we consider two metrics, Type-I (*α*) and Type-II (*β*) errors, which are defined by

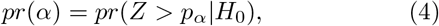

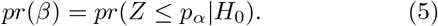

Here, Z is the estimator, *p*_*α*_ the critical bound and *H*_0_ the null hypothesis. We set the null hypothesis *H*_0_ to be the death of the patient in question, while the alternative hypothesis *H*_1_ to be the patient’s survival. Figure 4(a) shows the Type-I (*α*) and Type-II (*β*) errors from the hypothesis testing method for different stages of the treatment using the neural network outputs as estimators. Note that the highest accuracies (see Figure 4(b)) of each stage coincide with the accuracies obtained solely with the neural networks (see Table II).

**FIG. 4.**
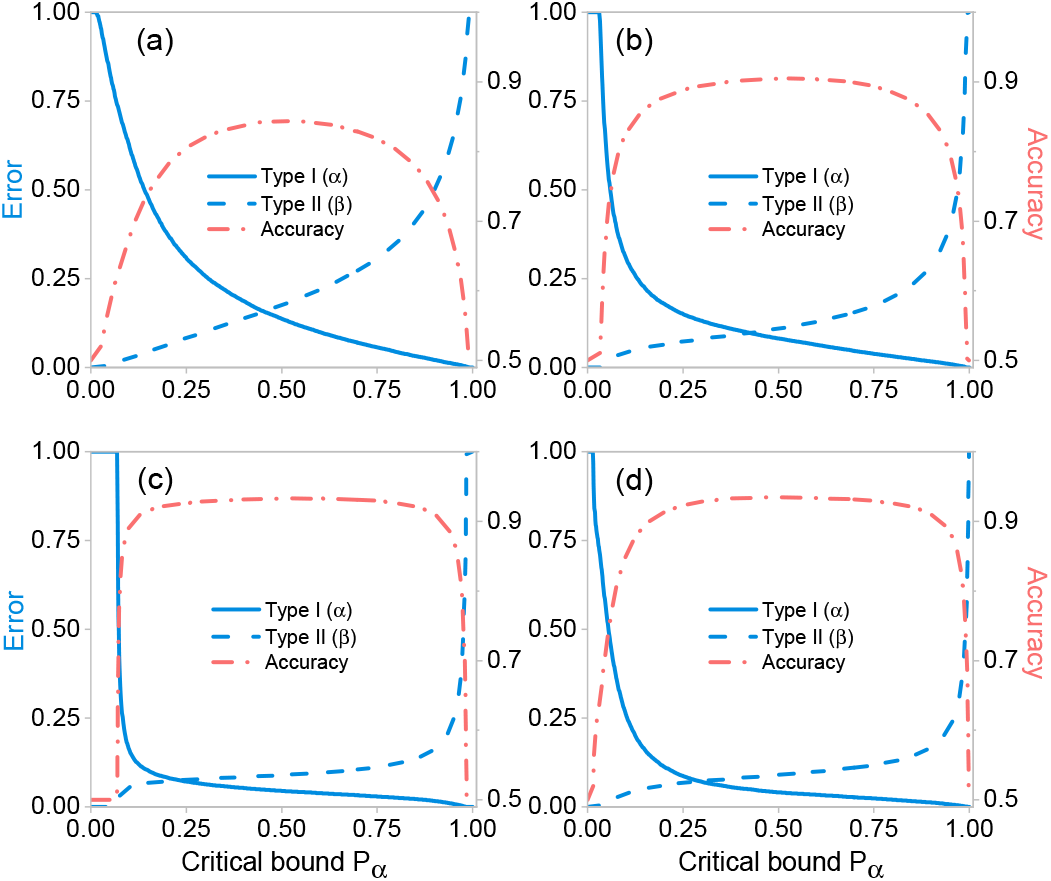
Type-I (*α*) and Type-II (*β*) errors, and accuracies for different treatment stages (a) Stage 1, (b) Stage 2, (c) Stage 3, and (d) Stage 4. Overall accuracy and errors are obtained from the hypothesis testing method as a function of critical bound *pα*, using the neural network outputs as estimators.

## VI. CONCLUSIONS

In the midst of this global crisis in which health care systems are overwhelmed, it is of utmost importance to focus efforts on the development of technological tools that allow achieving optimal use of health care resources. In this regard, it is essential to obtain significant knowledge of the prognostic factors associated with COVID-19 for its early identification. In this work, we have presented an effective machine-learning algorithm for the identification of high-risk patients, presenting COVID-19 symptoms. This technology enables rapid identification of high-risk patients at four different treatment stages, ranging from the onset of COVID-19–i.e. at the triage process for patients who arrive at the Emergency department–to the need for specialized care including and intensive care units. In order to train our neural networks, we have employed a characteristics vector with 21 elements per patient extracted from database which includes historical data for 4, 700, 464 confirmed or suspected COVID-19 cases. These 21 elements include information about comorbidities, demographical information, as well as recent, COVID-19-related medical information.

We have shown that our neural networks which contain two neurons in the hidden layer are capable of classifying with high accuracy patients into two classes: class 1, comprising those patients who are more likely to survive than to die, and class 2, comprising those patients who are more likely to die than to survive. Furthermore, we demonstrated that the accuracy, specificity, and sensitivity reach values up to 93.5%, 90.9%, and 96.1%, respectively.

Interestingly, we have shown that the training of our neural networks can accomplish the highly non-trivial task of determining an optimal estimator to be used as part of the standard hypothesis testing method. This results in a hybrid technique which can present the results of our artificial-intelligence-enabled patient classification algorithm in the language of hypothesis testing, commonly used in biostatistics and medicine, thus establishing a bridge between these two disciplines. We believe this result constitutes the foundation for a series of novel strategies for predicting outcomes in clinical medicine, and to enable new perspectives in clinical decision making. It is important to note that our algorithm could straightforwardly run on mobile phones or tablets, which could facilitate its deployment across clinics and hospitals. We point out that our research group plans to carry out prospective studies in which the tool presented here is to be applied to future COVID-19 patients as they seek medical attention, including public and private medical facilities with different budget levels located in urban areas with a range of population levels, thus allowing us to evaluate the ability of this new instrument to make useful predictions. We are certain that our work has important implications in the context of the current pandemic for medical resource allocation and hospital capacity planning.

## Data Availability

The database used in this work is publicly available in the Statistical Morbidity Yearbooks (Anuarios Estadisticos de Morbilidad) published by the General Council of Epidemiology (Direccion General de Epidemiologia), part of the Health Ministry (Secretaria de Salud), Mexican Federal Government.

https://www.gob.mx/salud/documentos/datos-abiertos-152127?fbclid=IwAR1V7f7WYhRn2DlDuPLJ7oGjL_0PR0XKdoInfUtf9DXJPj0dyVI0T7iz5kw

https://la.mathworks.com/matlabcentral/fileexchange/87202-idecovid19?s_tid=srchtitle

## ACKNOWLEDGMENTS

This work was supported by CONACyT under Project No. CB-2016-01/284372 and by DGAPA UNAM under Project UNAM-PAPIIT IN102920. A.U. acknowledges support from DGAPA-UNAM through grant UNAM-PAPIIT IN103521, and CONACyT through “Ciencia de Frontera” grant 217559.

## Appendix A Hypothesis testing with specific characteristics

As mentioned in Section III, the central moments computed from the characteristics vector for each patient do not constitute viable estimators to be used in hypothesis testing methods, as a consequence of the high degree of overlap between the resulting distributions for class 1 (recovered) and class 2 (deceased) patients. With this in mind, we have computed the central moments for a particular subset of characteristics found to be strongly correlated with the outcome (death/survival). Figure 5 shows the resulting distributions for the four first central moments using the following characteristics: age, hospitalization status, intubation, and ICU; class 1 (recovered) is shown in blue while class 2 (deceased) is shown in red. Clearly, the distributions are much less overlapped as compared to the case where all 21 characteristics are used (see Figure 1), allowing us to use these moments as viable estimators for hypothesis testing. Note, however, that the hospitalization status characteristic is only made available at stage 3 of treatment, while intubation and ITU at stage 4. Therefore, unfortunately, these estimators can only apply in an advanced treatment stages where the patients already are in need of specialized care.

**FIG. 5.**
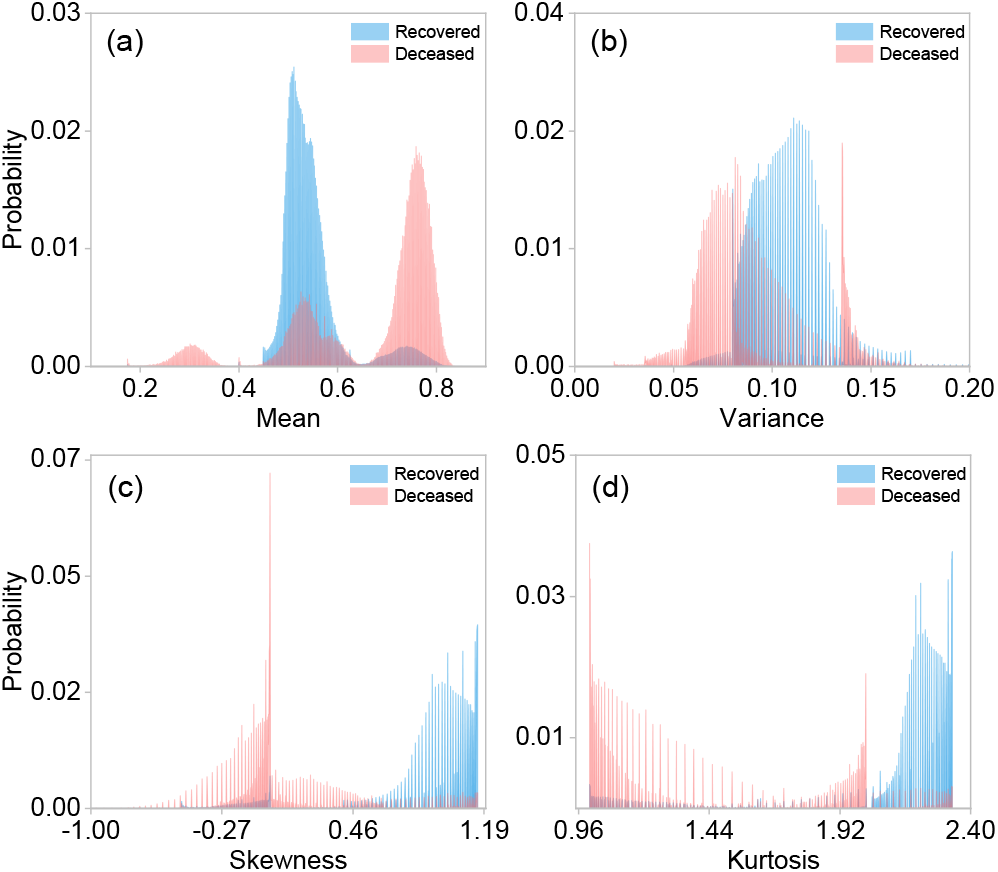
Normalized statistical distributions for different central moments: (a) Mean, (b) Variance, (c) Skewness, and (d) Kurtosis, for class 1 (recovered) shown in blue, and for class 2 (deceased) shown in red. These central moments were computed for a subset of 4 characteristics (age, hospitalization status, intubation and ICU). The distributions were computed from the whole dataset with 4, 700, 464 observations.

## Appendix B Confusion matrices and Receiver Operating Characteristic curves

In order to complement the results presented in this manuscript, we provide details about the performance of each of the trained artificial neural networks, for each of the four treatment stages, in terms of the confusion matrices, accuracies, and receiver operating characteristic curves (ROC). Figure 6 shows the confusion matrices which allow us to assess the performance of neural networks for the rapid identification of high-risk patients in each stage 1 through 4, presented in panels (a) through (d). Note that each matrix was computed from the 15% of observations reserved for testing. While the horizontal axis represents the known outcomes in the test dataset (target class), the vertical axis represents the predicted class using our neural networks (predicted class). Note that diagonal values represent successfully classified patients, i.e. true-positive and true-negatives, whereas off-diagonal elements represent misclassified patients, i.e. false-negatives and false-positives. Figure 6 also displays above each matrix the overall accuracy calculated from Eq. (3). Note that in all stages, the accuracy is larger than 84%.

**FIG. 6.**
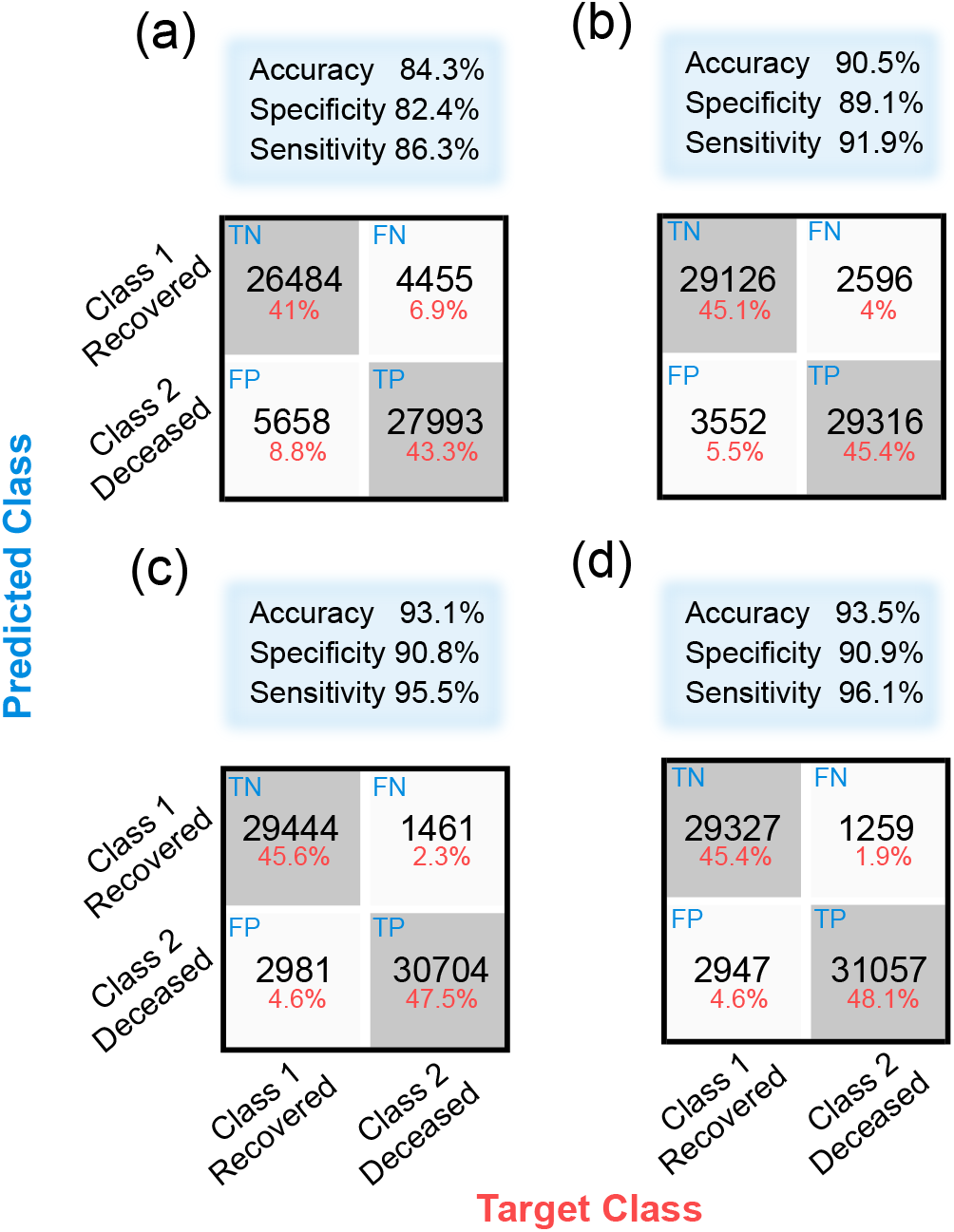
Confusion matrices, which quantify the performance of our neural networks in each of the four stages: (a) Stage 1, (b) Stage 2, (c) Stage 3, and (d) Stage 4. Off-diagonal values correspond to misidentified cases, i.e., false-negatives and false-positives, whereas the diagonal values represent true-positives and true-negatives.

Figure 7 shows the ROC curves of our neural networks for rapid identification of high-risk patients in each of the four treatment stages 1 through 4, shown in panels a through 4. These curves represent the sensitivity and specificity values computed across all possible threshold values. Here, the sensitivity is inversely related to the specificity, i.e., as sensitivity increases, the specificity decreases. Note that the curves shown in Fig. 7 show equivalent information to the ones presented in Section V for hypothesis testing methods in terms of the Type-I and Type-II errors.

**FIG. 7.**
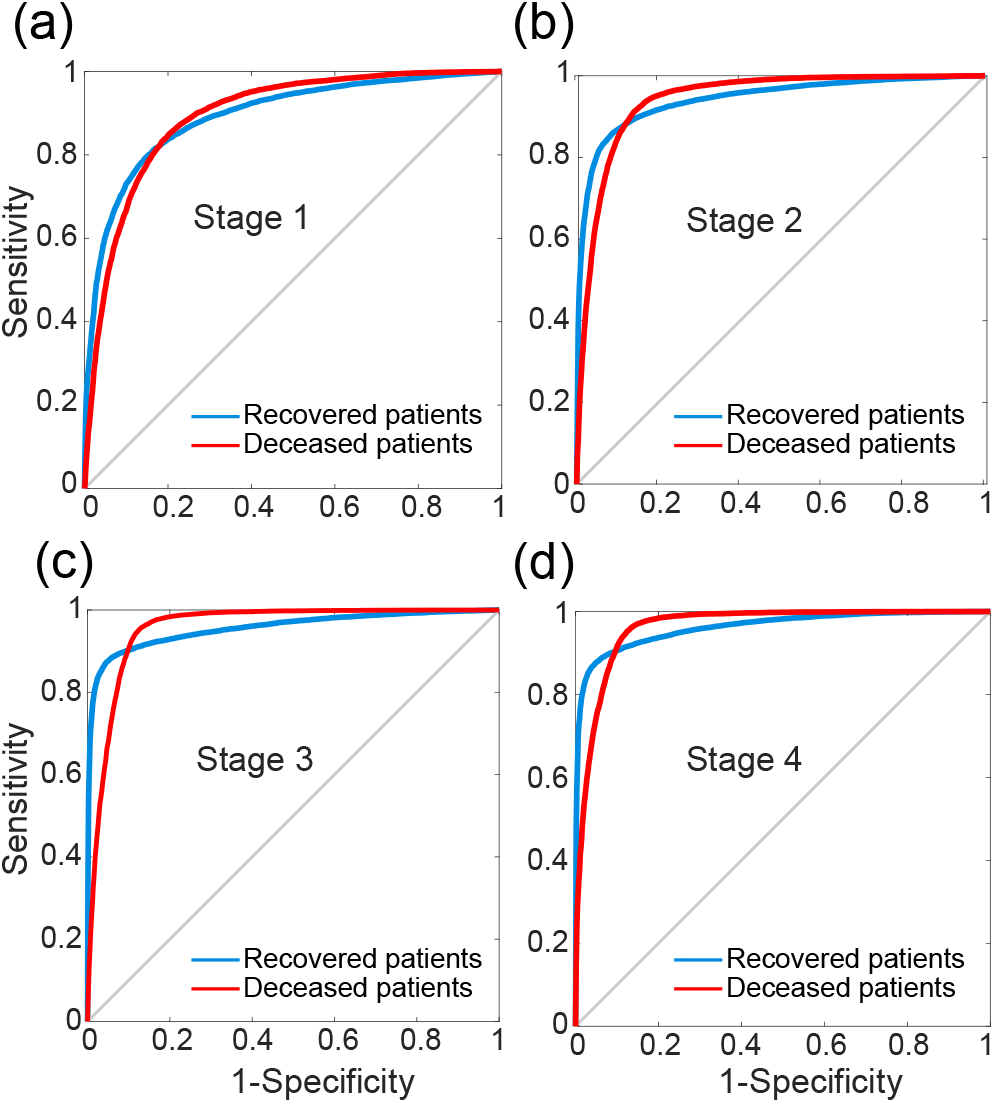
Receiver operating characteristic (ROC) curves that illustrate the diagnostic ability of the neural networks for rapid identification of high-risk patients in each stage: (a) Stage 1, (b) Stage 2, (c) Stage 3, and (d) Stage 4. The blue lines correspond to recovered patients, whereas the red lines depict the deceased patients.

Note that from our analysis it becomes possible to compare quantitatively the relative importance of each of the 21 characteristics in defining an outcome (death/survival) for a given patient. In Fig. 8 we plot the normalized, absolute value of the synaptic weights for neuron 1 (shown in blue) and neuron 2 (shown in red) as a function of the characteristic number (1 through 21, see table I), for each of the four treatment stages 1 through 4, shown in panels (a) through (d). We have also included from each of the two internal-layer neurons and for each of the four treatment stages, a list of the seven dominant characteristics, ranked by the absolute value of the synaptic weight (value shown within brackets).

**FIG. 8.**
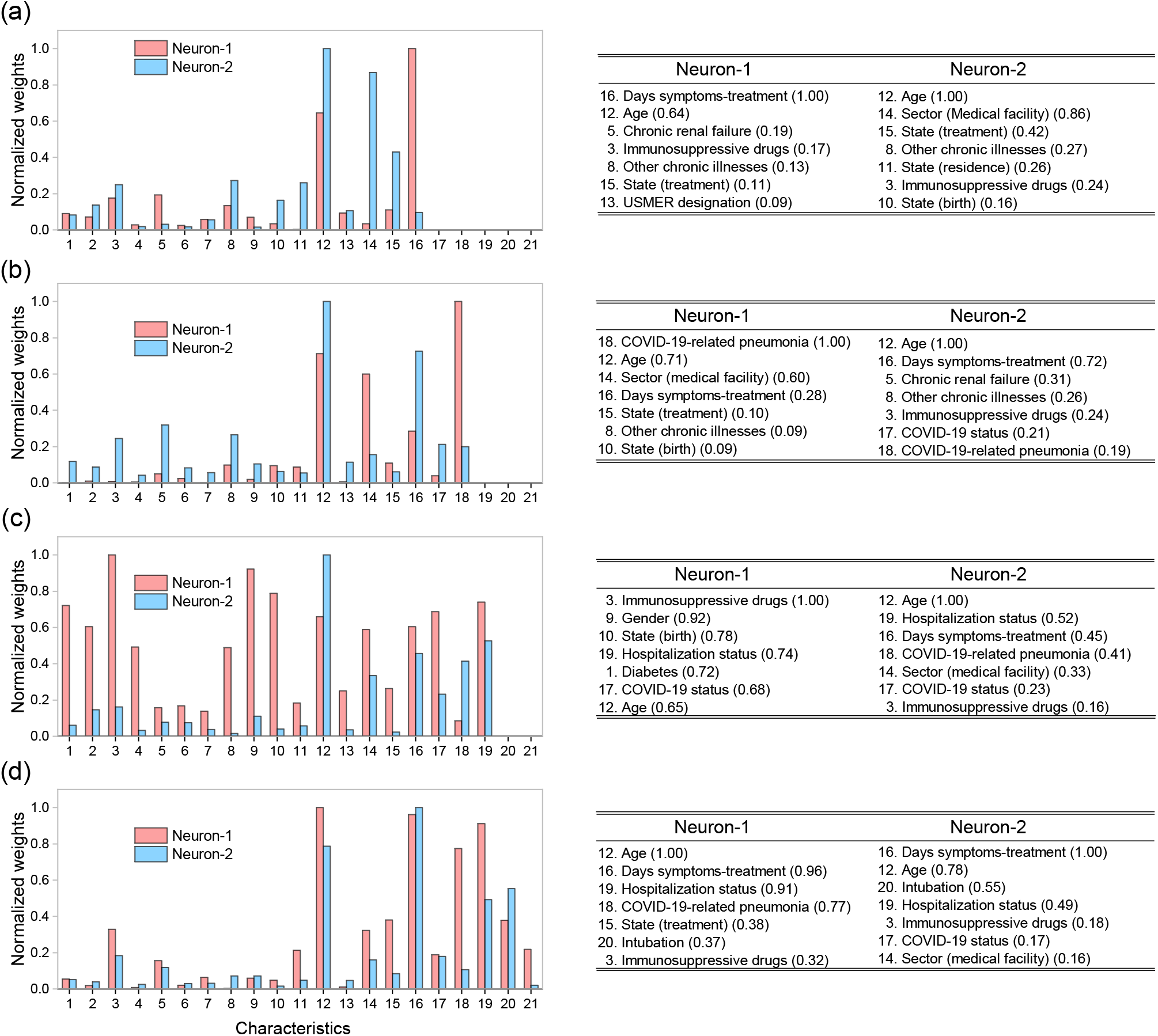
Left: Normalized absolute values of the synaptic weights for the two sigmoid neurons in the hidden layer for each stage: (a) Stage 1, (b) Stage 2, (c) Stage 3, and (d) Stage 4. Right: tables including the first seven dominant characteristics for each of the two neurons the hidden layer, ranked by the absolute value of the synaptic weight (value shown within brackets).

## Footnotes

1 including hospitals, clinics, and clinical laboratories

2 *USMER* is the acronym, in Spanish, used by the General Council of Epidemiology to appoint the health monitoring units of viral respiratory diseases (Unidades de Salud Monitoras de Enfermedad Respiratoria Viral) that integrate the Mexican epidemiological surveillance system. *Sector* refers to the type of institution, belonging to the National Health System, that provided the care.

